# Correction of Daily Positivity Rates for contribution of various test protocols being used in a pandemic

**DOI:** 10.1101/2020.08.25.20181347

**Authors:** Bhavik Bansal

## Abstract

Daily positivity rate (DPR) is a popular metric to judge the prevalence of an infection in the population and the testing response to it as a single number. It has been widely implicated in predicting future course of the SARS CoV-2 pandemic in India. With increasing use of multiple testing protocols with varying sensitivity and specificity in various proportions, the naïve calculation loses meaning particularly during comparison between states/countries with large daily variations in contribution of different testing protocols to the testing response. We propose an adjustment to the naïve DPR based on the testing parameters and the relative proportional use of each such protocol. Such a correction has become essential for comparing testing response of Indian states from Jun 2020 – Aug 2020 because of steep variations in testing protocol in certain states.

## Introduction & Background

The SARS Cov-2 pandemic in India has been generating all time high cases daily. Multiple parameters were introduced in wake of predicting the current course and predicting the future outcomes in various states. Daily Positivity Rates (DPRs) have been a go to metric that combines disease prevalence in an area with respect to testing adequacies. In addition to daily new cases, the daily test numbers were thought to represent a fuller picture, the logic being that additional testing would pick up infections with more sensitivity or correspondingly inadequate testing may under-reported positive cases. Their ratio is measured as DPR (Daily positivity rate) which we will call the ‘naïve DPR’ in this text.

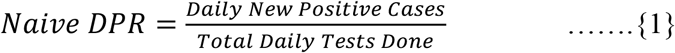

It was seen that when testing numbers were low, most of the cases that had received the test were symptomatic and therefore the fraction of tests that turn out positive were higher. This relationship between measured DPR and fraction of cases that are reported to the total number of people proposed to have been exposed (measured through sero-prevalence surveys) has been estimated to be a monotonically increasing polynomial of degree less than 1. [1]

It was also observed in some states (like Delhi) that DPR peaked before the new cases per day peaked and may therefore be a predictor of impending peak when corrected for confounders. (Figure 1)

**Figure 1.**
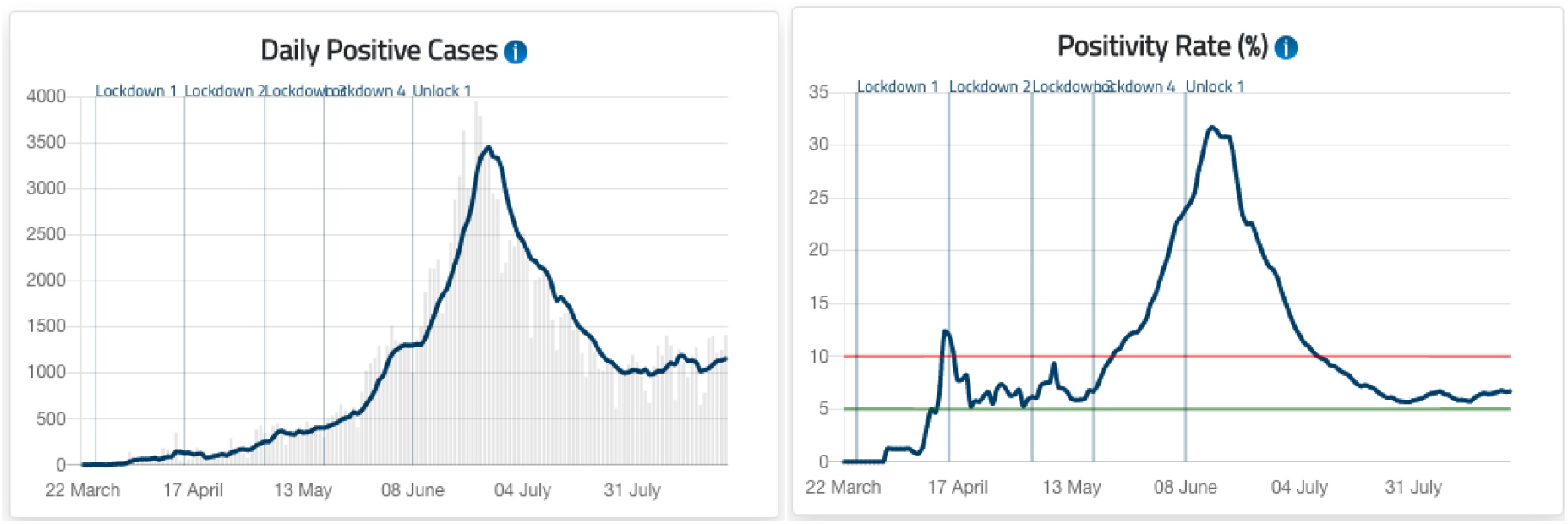
Both graphs are for Delhi – The blue lines in both cases are curves smoothed out by over 7 days weighing over the number of cases.(Illustrations from COVIDToday.in – an initiative by iCART – Indian COVID Apex Research Team)

## Problems with current methods

There are a multitude of tests available for SARS CoV – 2 infection detection in India, currently the RT-PCR (Reverse Transcriptase Polymerase Chain Reaction) is considered as the gold standard, while newer tests like the RAT (Rapid Antigen Tests) provide a faster though less sensitive (higher probability of a false negative) detection for the presence of an active infection.

With the advent of rapid antigen tests, which are believed to have ramped up testing in Delhi, there arose new ambiguity in the DPR metric. With other states like Karnataka, Andhra Pradesh and Kerala following suit by supplementing the RT-PCR (supposedly the gold standard) with other tests (with comparatively lower sensitivity scores), the naïve DPR metric does not stand indicative of what it was supposed to be, neither can it be compared to other states where RT-PCR is the exclusive test used. The sharp peak in Delhi has been often attributed to this bias by some sources.

Most states report testing number as a simple sum of all types of tests, which is believed to negatively bias DPR values because it treats all types of tests equivalent to the gold standard whereas the Rapid Antigen Test that has become a quite prevalent way to test for SARS CoV-2 can hardly be assumed so, with sensitivity score – (0.504,0.86) and specificity score – (0.993,1) reported for Standard Q COVID-19 Ag detection assay by SD Biosensor conducted independently by ICMR and AIIMS, New Delhi.[2]

## Data Availability

The basic reporting of new cases and total tests being done as a daily time series grouped by states has been available publicly for a long time now. Lately, some states have provided in addition to total testing metric, the fraction of tests done through various testing protocols. This fraction alongside the testing parameters (sensitivity and specificity) can be theoretically (with some assumptions and limitations) used to correct for this negative bias to some extent.

NOTE: Though this fractional data of testing is being provided by various state governments, their streamlined collection and analysis are missing right now. Volunteer team of data operators, where each volunteer spends about five minutes daily will enable the process. Each volunteer will be given a small instruction sheet, following which the volunteer has to enter data from a specified website onto a specified Google sheet, which will take less than five minutes daily. Multiple volunteers working like this in parallel on different data points will together enable daily complete data collection from all available sources. Our data science team will then work on pulling the data in a structured way and running analytical algorithms on it to provide daily analytics. The clean data sets will also be made available for public use to media, journalists and other researchers.

## Method

For given test parameters, to correct for the measured DPR we can use this expression:

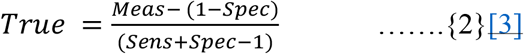

*True = Proposed true DPR in the sub — population that recieves the test*

*Meas = Sampled DPR through a given testing protocol*

*Sens, Spec = Test parameters for the test used for this sampling*

Inverting this function, we can evaluate ‘*Meas*’ by creating a function ‘*f*’of ‘ *True*’ DPR and test parameters (*Sens,Spec*), which samples a measured DPR for this test.

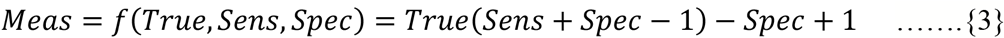

For any test whose testing parameters (sensitivity & specificity) are known, we can estimate the *‘Meas’* DPR as a function of the unknown *‘True’* DPR alone (here DPR may not necessarily mean prevalence in the population on a whole, rather a sub-population which gets tested, which cannot be extrapolated to the general population). As testing approaches infinity, DPR approaches true population prevalence.

Our algorithm fits the weighted average (based on the fraction of tests performed through a defined testing protocol) of the measured DPR (as a function of *’True’* DPR only, given the test parameters are known) for each test to the naïve DPR available from the daily tests and daily positive turnouts to estimate this *’True’* DPR (henceforth referred to as ‘corrected DPR’).

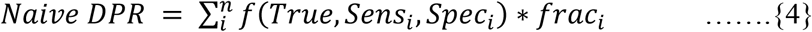

*Naïve DPR – as defined in eqn. {1}*

*True = Proposed true prevalance in the sub – population that recieves the test Sens_i_, Spec_i_ = Test parameters i^th^ testing protocol*

*frac_i_ = fraction of total tests done through the i^th^ protocol on a given day*

*n = number of testing protocols* (*here: 2 – RT-PCR and Rapid Antigen Tests*)

## Preliminary Results

Only a handful of states have reported this fractional testing data (not more than a couple month data at the maximum), not considering how inaccessible it is at this point. Though this data availability for testing distribution is not yet adequate, we hope to streamline this data and make it publicly available soon. Until then, we extracted the data for Karnataka (available from Jul 17 – Aug 6) and plotted naïve DPR (green) and corrected DPR (by above method in blue). We found that as the RAT became more abundant, the difference started to widen (Figure 2). Though we would discourage conclusions based on this dataset just yet because of the extremely small sample space.

**Figure 2.**
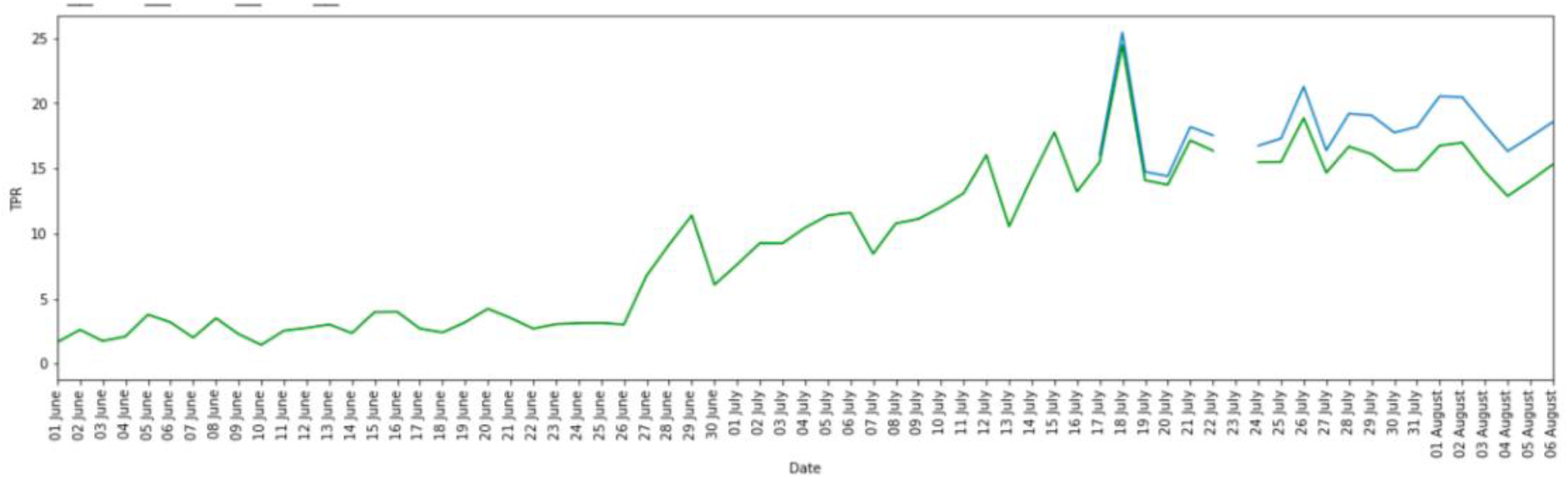
Original – DPR naïve (plotted in green) and DPR adjusted (plotted in blue) from daily (non-smoothed) dataset by method illustrated above. This considers RT-PCR as gold standard and RATs having sensitivity of 0.7 and specificity of 0.996.

## Discussion

This kind of correction is best applicable when there exist sharp changes in the ratios of testing on a daily bases and among comparator states/countries, which basically describes Indian states in Jun 2020 – Aug 2020. Data already suggests large variation of testing parameters and the measured positivity rate with different labs, the fraction of contribution each lab/protocol makes in the net positivity rate is a major issue.

This method can be extended for DPR correction over more than 2 types of tests with adequate data on each test parameters. Therefore it can be extended to diseases other than SARS CoV-2 wherein multiple diagnostic tests with varying sensitivities/specificities are to be combined into a single parameter of significance.

This method can therefore be further extended to take into account differing test parameters for different kits of the similar test architecture (various kinds of RAT will be soon be available) given adequate information of their independent testing parameters is available.

Also unlike other sensitivity based corrections, this method doesn’t assume any test parameter to be fixed, one can vary both sensitivity and specificity (even for RT-PCR which has been traditionally regarded as both values = 1 i.e. defined as the gold standard) which can be of use if in the future a better gold standard is proposed. [4],

### The Major Assumption

The underlying assumption is that the hypothetical True DPR is same for all testing protocols, a much lighter one than assuming similar measured DPR for varying protocols as was done previously. But it is not completely justified to assume equal prior probabilities for positivity for tests as different as RT-PCR and RAT. But we discuss later using data from Delhi and Karnataka, that for these states, this assumption is not as baseless as one might believe.

The RT-PCR (and equivalent tests like CBNAAT & TrueNat) and the Rapid Antigen tests are not exactly testing the same population. It is evident through data as well as reported by experts on front lines that the sub-set of population that receives the respective tests are anything but similar. Generally, more severe cases are supposed to receive an RT-PCR while lesser ones to receive a RAT therefore the assumption of a common ‘True DPR’ for the tests is questionable.

The recent ICMR guidelines also recommend an RT-PCR as a required follow-up when an initial negative Rapid Antigen Test has been performed whereas a positive RAT be considered a true positive (relying on its high specificity). [2] If the fraction of RT-PCRs done for such follow-ups constitute a significant fraction of the total RT-PCRs, our initial assumption of both tests having similar True DPR would fail miserably.

To test this, we obtained data on RATs done and their results with follow-ups during Jun 18 – Jul 15 2020 in various districts in Delhi. [5]

It was found that among the total tests (n = 415,134) done in Delhi during this period, RATs (n = 281,555) constitute 67.7% of them, of whom at net 6.92% turned out positive (this is the measured DPR for RATs). Of the RATs that turned out negative (n = 262,075), only a meagre 0.52% (n = 1,365) were followed-up with a RT-PCR.

Similar analysis for the total RT-PCRs (n = 133,579) of whom 47,291 turned out positive, gave a net positivity rate (here: measured) of 35.40%.

The fraction of PCRs that were employed as a follow-up to negative RATs was only 1.02%, which gives certain validity to our initial assumption and dilutes the extent of the limitation. From the data of PCR follow-ups, extrapolating to all RAT negative cases, we also managed to estimate an independent measure of sensitivity of RAT (assuming their specificity is almost 1). This estimated yielded an average sensitivity of 0.29, varying from 0.06 to 0.59 among districts.

The extrapolation of RAT False Negatives was done as:

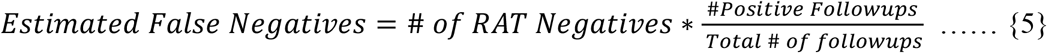

The sensitivity of RAT was estimated as follows:

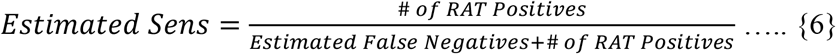

When corrected for measured sensitivity of RAT (assuming its specificity = 1), the true positivity can be evaluated to be around 23.86%.

**Table 1:** District wise RAT follow-up data from Delhi (18 Jun – 15 July 2020) [5]

| District | Total RAT<br>(18 Jun -<br>15 July) | # of RAT<br>Positive | # of RAT<br>Negative | % of RAT<br>Positive | # of RT-PCR<br>follow up | # of<br>positive<br>follow ups | % of<br>follow<br>up | Estimated<br>Sensitivity of<br>RAT |
| --- | --- | --- | --- | --- | --- | --- | --- | --- |
| South | 16421 | 2137 | 14284 | 13.01 | 97 | 10 | 0.68 | 0.59 |
| South<br>West | 39804 | 3798 | 36006 | 9.54 | 38 | 11 | 0.11 | 0.27 |
| Shahdara | 28009 | 2571 | 25438 | 9.18 | 648 | 123 | 2.55 | 0.35 |
| South<br>East | 23221 | 2073 | 21148 | 8.93 | 37 | 9 | 0.17 | 0.29 |
| West | 27692 | 2048 | 25644 | 7.40 | 123 | 15 | 0.48 | 0.40 |
| East | 22003 | 1289 | 20714 | 5.86 | 180 | 24 | 0.87 | 0.32 |
| North<br>West | 17604 | 930 | 16674 | 5.28 | 48 | 9 | 0.29 | 0.23 |
| New<br>Delhi | 23979 | 1127 | 22852 | 4.70 | 60 | 8 | 0.26 | 0.27 |
| Central | 25561 | 1167 | 24394 | 4.57 | 86 | 6 | 0.35 | 0.41 |
| North<br>East | 24929 | 1052 | 23877 | 4.22 | 28 | 18 | 0.12 | 0.06 |
| North | 32332 | 1288 | 31044 | 3.98 | 20 | 10 | 0.06 | 0.08 |
| Total | 281555 | 19480 | 262075 | 6.92 | 1365 | 243 | 0.52 | 0.29 |

Similar data was available for Karnataka from Jun 25 to Aug 5 2020 [6], which revealed a similar story of dismal RT-PCR follow-ups to negative RATs. It was found that among the total tests (n = 979,329) done in Karnataka during this period, RATs (n = 290,085) constitute 29.6% of them, of whom at net 12.87% turned out positive (this is the measured DPR for RATs). Of the RATs that turned out negative (n = 252,726), only a meagre 5.6% (n = 14,209) were followed-up with an RT-PCR, though better than Delhi but not significant enough to come out as a major limitation.

Similar analysis for the total RT-PCRs (n = 689,244) of whom 140,889 turned out positive, gave a net positivity rate (here: measured) of 20.44%.

Similar sensitivity was estimated (as in equations {5} and {6}) to yield an estimate of 0.43. Both these estimated sensitivities are lower than the ICMR study supporting the need of such a correction to naïve DPR as proposed.[2]

When corrected for measured sensitivity of RAT (assuming its specificity = 1), the true positivity can be evaluated to be around 29.93%.

Hence, in the cases of Delhi and Karnataka from the data available publicly, it can be seen that True DPR of RAT and RT-PCR are indeed not as different as would be predicted given all protocols were religiously followed, therefore our correction is at least justified for the Indian context from Jun 2020 – Aug 2020.

## Conclusion

The proposed corrected DPR takes into account varying test parameters and fractional abundance assuming same prior prevalence. This correction can significantly improve interpretation and comparison of prevalence and testing data between states through time during the growth phase of a pandemic, particularly when the steep variations in testing distribution is seen.

## Data Availability

All data referred to in the manuscript are publicly accessible and were released by state governments in daily bulletins. Most of the data mined data was procured from COVIDToday.in

https://covidtoday.in/

